# Deep Learning-Based Opportunistic CT Osteoporosis Screening and Establishment of Normative Values

**DOI:** 10.1101/2025.03.25.25324606

**Authors:** Malte Westerhoff, Soterios Gyftopoulos, Bari Dane, Emilio Vega, Daniel Murdock, Norbert Lindow, Felix Herter, Khaled Bousabarah, Michael P. Recht, Miriam A. Bredella

**Affiliations:** Visage Imaging, GmbH, Berlin, Germany; Department of Radiology, NYU Langone Health and NYU Grossman School of Medicine, New York, NY, USA

## Abstract

**Background:** Osteoporosis is underdiagnosed and undertreated prompting the exploration of opportunistic screening using CT and artificial intelligence (AI).

**Purpose:** To develop a reproducible deep learning-based convolutional neural network to automatically place a 3D region of interest (ROI) in trabecular bone, develop a correction method to normalize attenuation across different CT protocols or and scanner models, and to establish thresholds for osteoporosis in a large diverse population.

**Methods:** A deep learning-based method was developed to automatically quantify trabecular attenuation using a 3D ROI of the thoracic and lumbar spine on chest, abdomen, or spine CTs, adjusted for different tube voltages and scanner models. Normative values, thresholds for osteoporosis of trabecular attenuation of the spine were established across a diverse population, stratified by age, sex, race, and ethnicity using reported prevalence of osteoporosis by the WHO.

**Results:** 538,946 CT examinations from 283,499 patients (mean age 65 years±15, 51.2% women and 55.5% White), performed on 50 scanner models using six different tube voltages were analyzed.

Hounsfield Units at 80 kVp versus 120 kVp differed by 23%, and different scanner models resulted in differences of values by < 10%. Automated ROI placement of 1496 vertebra was validated by manual radiologist review, demonstrating >99% agreement. Mean trabecular attenuation was higher in young women (<50 years) than young men (p<.001) and decreased with age, with a steeper decline in postmenopausal women. In patients older than 50 years, trabecular attention was higher in males than females (p<.001). Trabecular attenuation was highest in Blacks, followed by Asians and lowest in Whites (p<.001). The threshold for L1 in diagnosing osteoporosis was 80 HU.

**Conclusion:** Deep learning-based automated opportunistic osteoporosis screening can identify patients with low bone mineral density that undergo CT scans for clinical purposes on different scanners and protocols.

**Key Results 3 main results/conclusions:**

- In a study of 538,946 CT examinations performed in 283,499 patients using different scanner models and imaging protocols, an automated deep learning-based convolutional neural network was able to accurately place a three-dimensional regions of interest within thoracic and lumbar vertebra to measure trabecular attenuation.
- Tube voltage had a larger influence on attenuation values (23%) than scanner model (<10%).
- A threshold of 80 HU was identified for L1 to diagnose osteoporosis using an automated three-dimensional region of interest.

## Introduction

Osteoporosis is characterized by low bone mineral density (BMD), which increases the risk of fractures (1). Even though osteoporosis is both preventable and treatable, it remains underdiagnosed and undertreated (2). Consequently, screening guidelines have been established to identify reduced BMD in at-risk individuals (3). Among patients eligible for osteoporosis screening, only one out of five patients undergo screening (4). In addition, significant disparities exist in osteoporosis screening and treatment across different racial and ethnic groups (5, 6).

Osteoporosis screening is typically performed by dual-energy x-ray absorptiometry (DXA), however, 50% of those who suffer a fracture have BMD values on DXA in the normal range (7, 8). BMD assessed by DXA depends on body size, which can lead to erroneous assessments of fracture risk for small patients (9) and those with obesity (10). DXA values in the spine can also be impacted by degenerative changes, such as osteophytes or vascular calcifications, which are common in the elderly.

This can falsely elevate BMD and underestimate fracture risk (9). Moreover, DXA is a 2D planar technique that cannot separate cortical from trabecular bone (11).

Quantitative CT (QCT) using a calibration phantom is a more accurate method for measuring BMD compared to DXA (12). QCT is able to measure volumetric BMD, separate cortical from trabecular bone, and is less susceptible to overestimations of BMD due to degenerative diseases (11, 13). More recently, an “opportunistic screening” approach with AI-driven automatic assessment of BMD of the spine has been proposed to detect osteoporosis of the spine using non-contrast CT scans performed for other clinical purposes (14, 15). Potential sources of inaccuracy with CT evaluation of BMD include variation in trabecular attenuation within a vertebra when using a 2D region of interest (ROI) on a single axial slice (14), and differences in scan parameters, such as difference in tube voltages, or scanner model (16, 17). There is also a lack of established normal values for BMD by opportunistic CT in racially and ethnically diverse populations. This can influence the performance of risk prediction algorithms and possibly lead to unequal outcomes for individuals from diverse backgrounds (18, 19).

The purpose of this study was threefold. First, we aimed to develop a reproducible and robust deep learning-based method using a convolutional neural network (CNN) that can automatically and accurately place a 3D ROI in trabecular bone. Second, we aimed to develop a statistical correction method to normalize attenuation of the thoracic and lumbar spine across different CT protocols and scanner models. Third, we aimed to establish normative values for CT measurements of spine trabecular attenuation in a large diverse population over a wide age range.

## Materials and Methods

### Patient population

This study was approved by the institutional review board and was Health Insurance Portability and Accountability Act compliant. The requirement for informed consent was waived. A retrospective picture archiving and communication system (PACS; Visage Imaging, Inc., San Diego, CA, United States) search was performed for all patients between 20 and 94 years of age who underwent a non-dual energy, non-contrast CT of the chest, abdomen, thoracic or lumbar spine between 2003-2024 with a slice thickness of at least 5mm. The electronic health record was reviewed to obtain self-reported race and ethnicity information. As this study aimed to screen patients who would be diagnosed with osteoporosis or osteopenia by DXA of the spine or hip, patients on medication that could affect bone metabolism were included, similar to the National Health and Nutrition Examination Survey (NHANES), which serves as the reference database for DXA (20).

### Computed Tomography Technique

Non-contrast chest, abdominal, and spine CT examinations were performed according to standard departmental protocols at 80-130 kVp. Tube current was 50 to 160 mAs, pitch 0.6-0.9, and rotation time 0.25-0.625s. Axial, coronal, and sagittal images at 0.5-5 mm were generated.

### Automated Image segmentation

To automate ROI placement, a CNN was trained to segment the CT volumes into the major anatomical structures, specifically including and labeling the vertebral bodies as described by *Wasserthal et al* (21). The standard method for manual measurement of bone attenuation on CT images typically involves placing a circular ROI within a single axial slice of the trabecular bone of a suitable vertebral body and measuring the mean attenuation in Hounsfield Units (HU). The choice of slice and ROI positioning can significantly impact the result because of trabecular inhomogeneities within a vertebral body (14). We therefore used 3D spherical ROIs, which cover more voxels and therefore are less susceptible to trabecular inhomogeneities.

The volume of each lumbar and thoracic vertebra that was fully contained within the field of view was automatically calculated and a spherical ROI with a volume of 5% of the respective vertebral body was placed inside the vertebra. The ROI position was determined automatically to meet the following criteria: (i) the entire ROI was fully contained within the respective segmented vertebra, (ii) it had the lowest attenuation (14), and (iii) it was placed anterior to the basivertebral foramen **(Figure 1)**. The median HU within the spherical ROI was then computed to provide a more robust measure against outliers than could be seen using a mean value. Finally, the HU values were averaged for each vertebral body that was fully imaged in more than one series.

**Figure 1.**
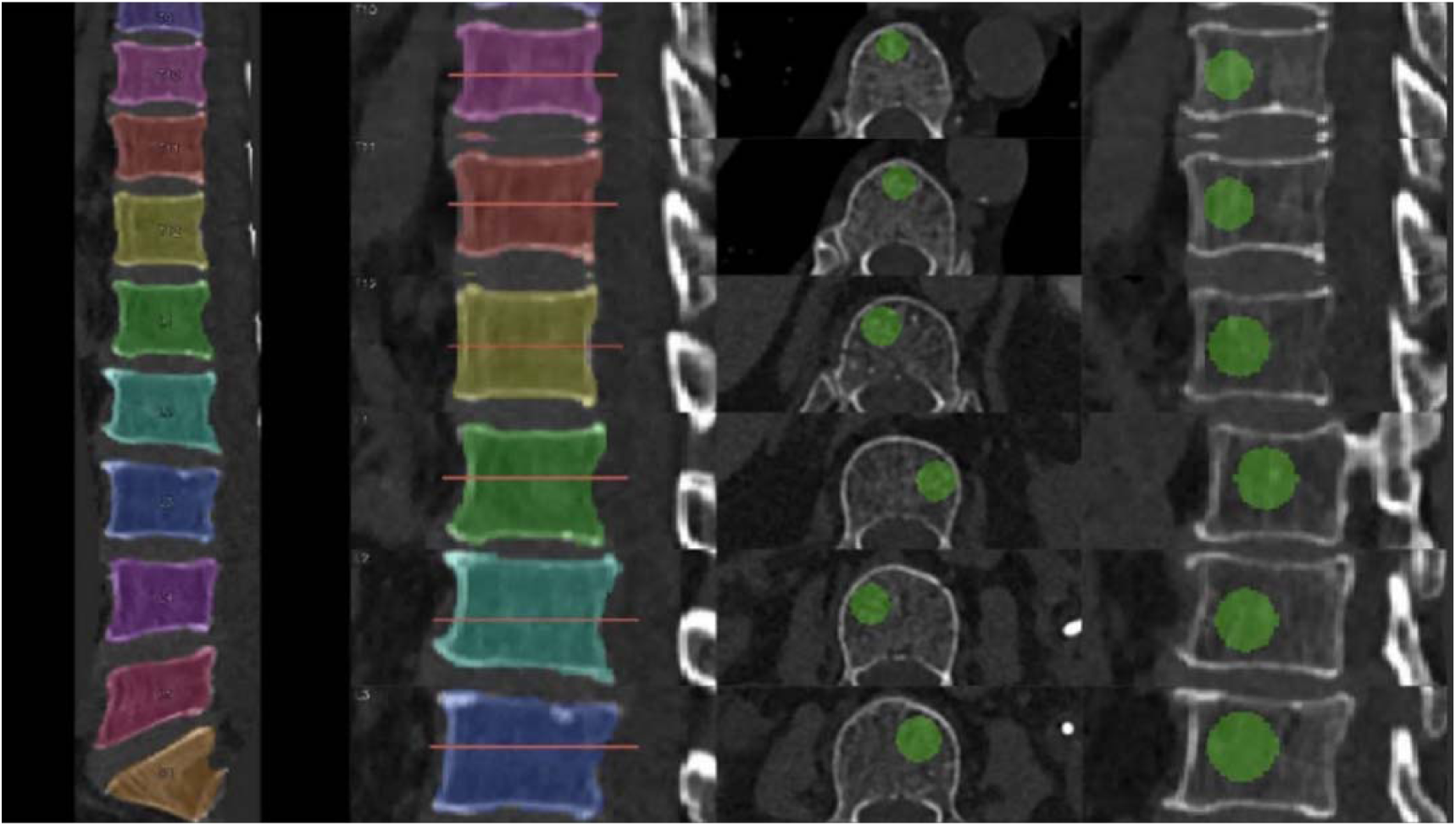
Placement of 3-dimensional region of interests within thoracic and lumbar vertebral bodies. a) straightened spine (curved view) with vertebrae segmentation, b) close-up of vertebra segmentation with axial ROI center slice position indicator, c) axial ROI placement, and d) sagittal ROI placement.

Automated consistency checks for each vertebral body were performed to ensure that the segmentations were valid **(Supplement 1)**. If they were not valid, the series was excluded Three board certified musculoskeletal radiologists with ≥20 years of experience (SG, MPR, MAB) independently reviewed segmentations and ROI placements in1496 lumbar and thoracic vertebrae in 128 CT scans from 128 patients randomly selected that were not part of the training set.

To compare attenuation values obtained by our 3D method with the standard 2D method, we also automatically placed a 2D elliptic ROI in the central slice of each vertebral body and calculated the mean attenuation as described by Jang et al. (14).

### Adjustments for Tube Voltage and Scanner Model

Attenuation values in HU from different scanner models and different kVp values were calibrated to a common reference, as the included CT examinations were performed on multiple different CT scanner models with different imaging protocols **(Supplement 2)**.

The most commonly used tube voltage for the included studies was 120 kVp (60% of studies) and thus served as the reference for our data. For scans with other tube voltages, the HU values were adjusted to the equivalent 120 kVp using a linear scaling HU_120_ = A_x_ HU_x_ where HU_x_ is the HU measured at voltage x, HU_120_ is the equivalent value calibrated to a 120 kVp scan, and A_x_ is the mapping parameter determined by the statistical analysis from **Supplement 2**. After tube voltage calibration, scanner model calibration was performed using the same statistical method **(Supplement 2)**.

### Statistical analysis

Statistical analysis was performed using software (SQL Server and Excel, Microsoft, Redmond, WA, and Python 3.12). Continuous variables were reported as mean, median, standard deviation, 10^th^ and 90^th^ percentiles. Categorical variables were reported using counts and percentages. Normative values for each lumbar and thoracic vertebral body were stratified by sex, race, and ethnicity, and age group in 5-year intervals. We used all eligible series of each CT to maximize the statistical power of our analysis. To avoid over-representing studies with many series, or patients with many studies, we first calculated the mean value for each vertebra across all series in each study. We then calculated the mean value for all scans from the same patient that fall within the same age group. Using these mean values, we then determined the trabecular attenuation distribution for each vertebra, sex, and age group. Differences between groups were assessed using the Student’s t-test. A two sided p value <.05 was considered significant.

The standard criterion to diagnose osteoporosis by DXA is a BMD that is-2.5 standard deviations (SD) or lower than that of a healthy young population (T score-≤2.5), and the diagnosis of osteopenia is a BMD that is between-1 and-2.5 SD below the BMD of a healthy young population (22). Given that our proposed method is a different measurement technique than DXA, statistical measures such as standard deviation are different and therefore the same thresholds cannot be applied (23). To establish thresholds that correspond to the established DXA criteria of osteoporosis, osteopenia, or normal, we used the reported prevalence of 22% of osteoporosis in 60-69 year-old female patients as determined by the WHO (24). The prevalence of low bone mass, i.e. osteopenia or osteoporosis, has been estimated to be 65 % (25). Consequently, the 22^nd^ (24) and 65^th^(25) percentiles of CT-based attenuation (HU) were calculated for each vertebra in our female patient population. The International Society for Clinical Densitometry (ISCD) recommended that a female reference database should be used to assess osteoporosis in men (22). We therefore did not determine separate thresholds for men.

In order to assess whether our sample of patients who underwent CT is representative of the general population with regards to their statistical bone density distribution, we compared osteoporosis prevalence among all women aged 60–69 undergoing DXA screening at our institution with the subset that also had a CT within 12 months of the DXA.

## Results

### Demographics

In our retrospective PACS search 592,878 non-contrast CT examinations in 307,238 patients were identified. The most common indications were renal (suspected kidney stone), pulmonary (nodules, cough, pneumonia, bronchiectasis), back pain, and trauma. Based on the automated consistency checks, 53,932 CTs of 23,739 patients were excluded due to technical reasons such as metal artifact, compression fractures, sclerotic or lytic vertebral lesions that precluded accurate assessment of trabecular attenuation, resulting in 538,946 evaluable CT examinations in 283,499 patients (mean age 65 years ± 15, 51.2 % women and 55.5 % White) **(Figure 2 and Table 1).**

**Figure 2.**
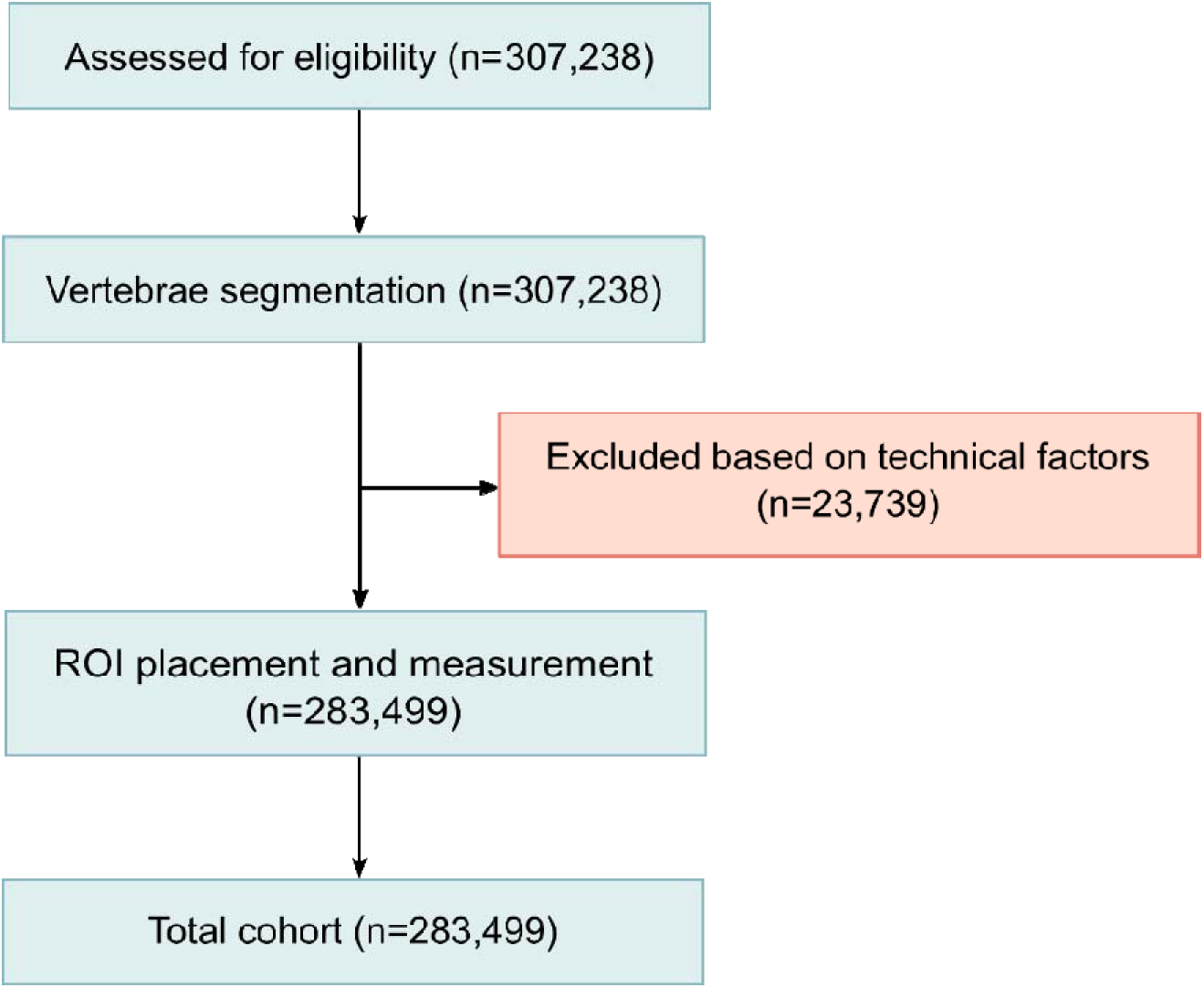
Flow diagram

**Table 1.** Demographics of the study population.

| Variables | Mean (SD) |
| --- | --- |
| Age (years) | 65 ± 15 |
| Female (%) | 145,021 (51.2%) |
| Male (%) | 138,478 (48.8%) |
| Race (%) |  |
| White | 157,457 (55.5%) |
| Black | 24,152 (8.5%) |
| Asian | 13,938 (4.9%) |
| Other/unknown | 87,952 (31.0%) |
| Ethnicity (%) |  |
| Hispanic | 22,114 (7.8%) |
| Non-Hispanic | 152,596 (53.8%) |
| Other/unknown | 108,789 (38.4%) |

### Automated ROI placement, Scanner and kVp Model Adjustments

In 99.9% (1495 of 1496 vertebra), the radiologists agreed with the automated ROI placement.

The mean difference between 2D vs 3D ROI placement was 53HU and the median difference was 26 HU (14-26 IQR).

CTs were performed on 50 scanner models using six different tube voltages. Mapping parameters (Ax) for each tube voltage (kVp) to adjust HU to the equivalent of 120 kVp, the most commonly used tube voltage, are listed in **Table 2a**. HU at 80 kVp and 120 kVp differed by 23% (correction factor 0.77).

**Table 2a.** Adjustment of Hounsfield Units (HU) values to the equivalent 120 kVp using linear scaling.

| **Tube Voltage (kVp)** | **Mapping parameter Ax** |
| --- | --- |
| 80 | 0.773 |
| 90 | 0.808 |
| 100 | 0.918 |
| 110 | 0.935 |
| 120 | 1.000 |
| 130 | 1.015 |

**Table 2b.**
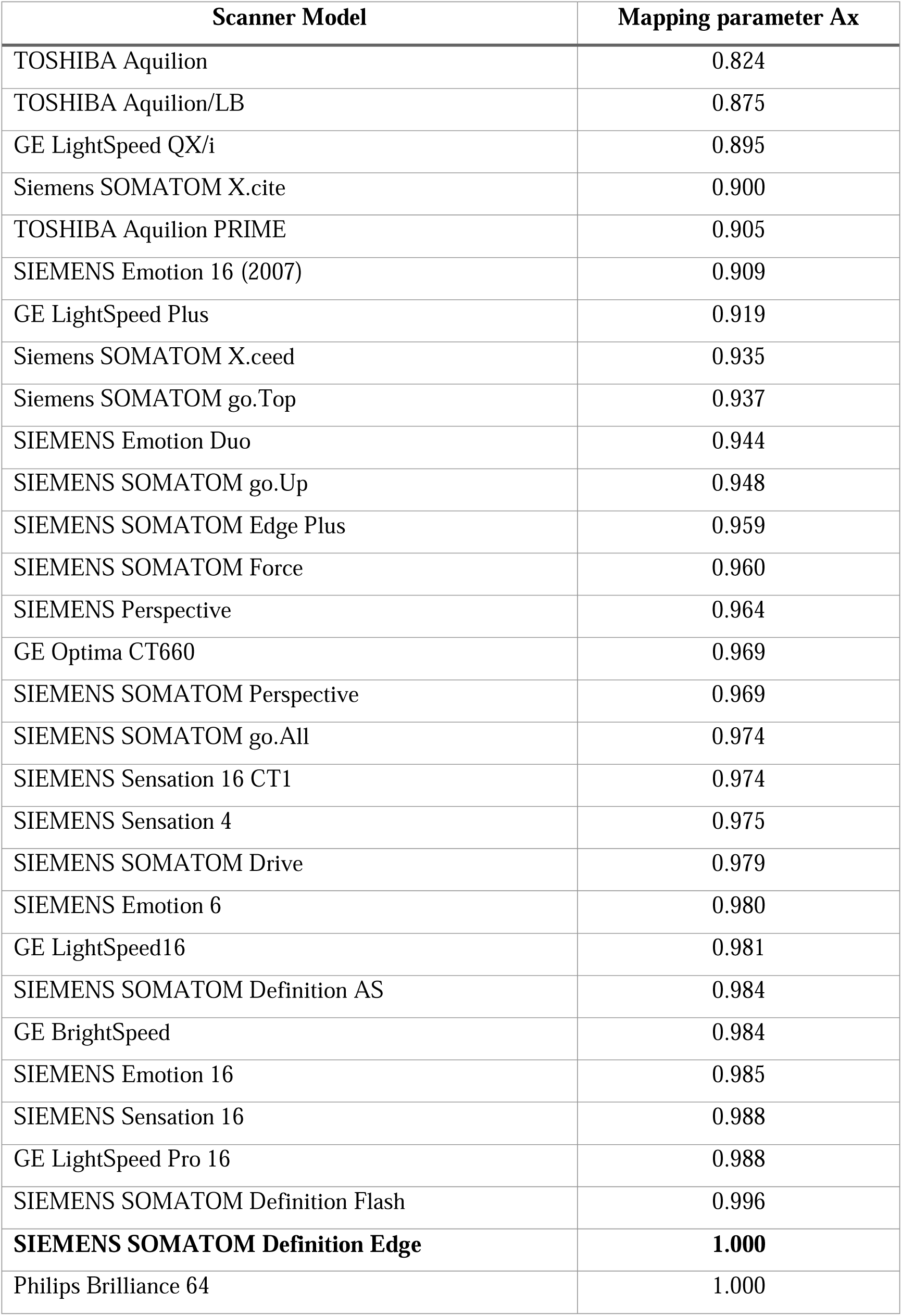
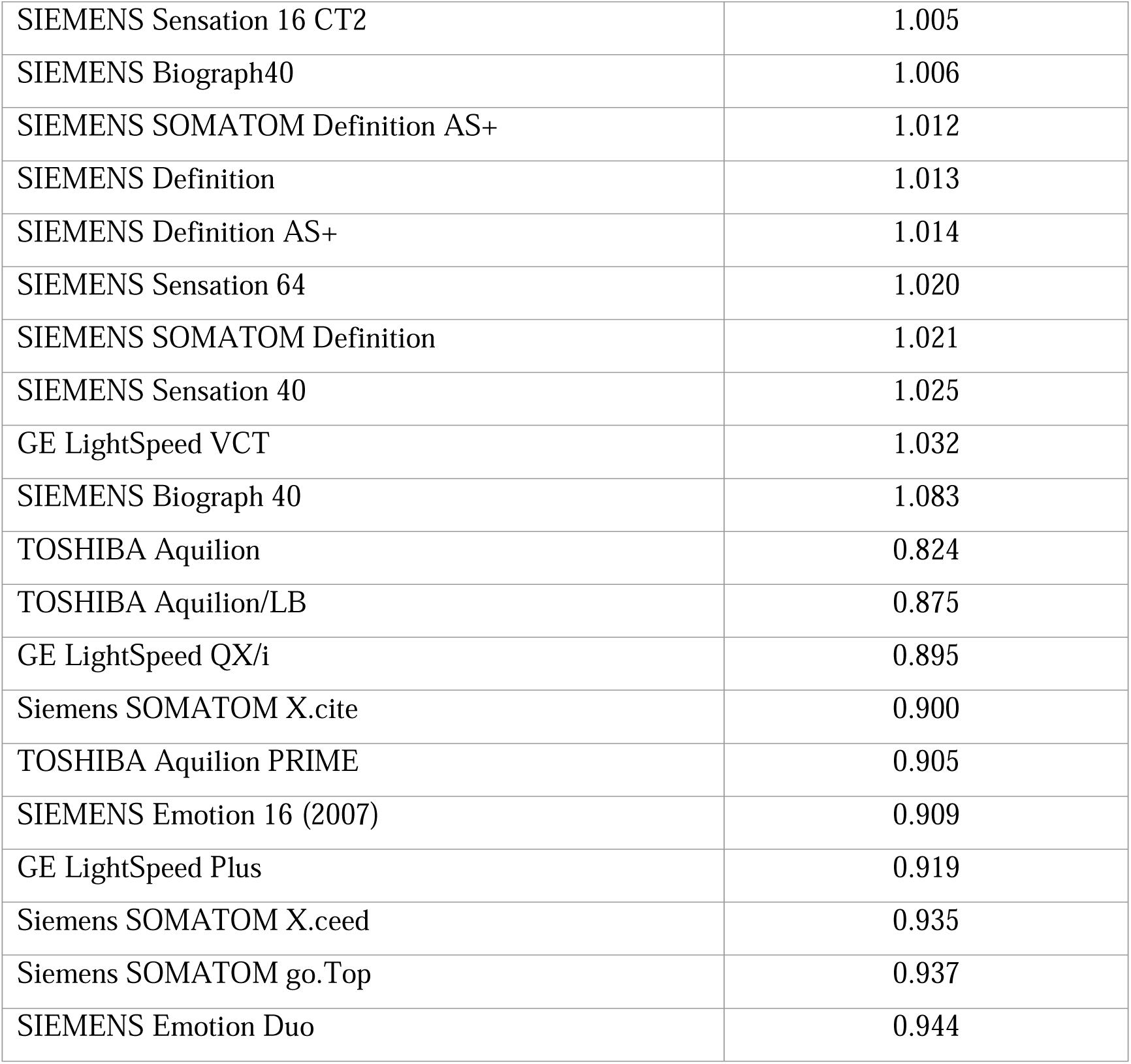
Scanner models and mapping parameter using linear scaling.

**Table 3a.** Normative attenuation values of L1 trabecular bone in Hounsfield Units (HU) in females.

| **Age (years)** | **Mean** | **10<sup>th</sup> percentile** | **Median** | **90<sup>th</sup> percentile** |
| --- | --- | --- | --- | --- |
| 20 - 24 | 213 | 166 | 213 | 262 |
| 25 - 29 | 207 | 157 | 206 | 259 |
| 30 -34 | 204 | 154 | 202 | 257 |
| 35 -39 | 200 | 149 | 198 | 254 |
| 40 - 44 | 191 | 139 | 189 | 246 |
| 45 - 49 | 177 | 123 | 176 | 233 |
| 50 - 52 | 152 | 100 | 148 | 208 |
| 55 -59 | 130 | 84 | 127 | 180 |
| 60 -64 | 116 | 71 | 112 | 165 |
| 65 - 69 | 105 | 61 | 102 | 153 |
| 70 -74 | 97 | 53 | 93 | 144 |
| 75 - 79 | 88 | 44 | 84 | 135 |
| 80 - 84 | 81 | 36 | 76 | 127 |
| 85 - 89 | 73 | 28 | 68 | 120 |
| 90 -95 | 66 | 20 | 62 | 114 |

**Table 3b.** Normative attenuation values of L1 trabecular bone in Hounsfield Units (HU) in men.

| **Age (years)** | **Mean** | **10<sup>th</sup> percentile** | **Median** | **90<sup>th</sup> percentile** |
| --- | --- | --- | --- | --- |
| 20 - 24 | 206 | 156 | 204 | 261 |
| 25 - 29 | 199 | 149 | 197 | 251 |
| 30 -34 | 190 | 142 | 189 | 241 |
| 35 -39 | 183 | 134 | 180 | 234 |
| 40 - 44 | 173 | 124 | 171 | 224 |
| 45 - 49 | 164 | 115 | 161 | 215 |
| 50 - 52 | 151 | 103 | 148 | 204 |
| 55 -59 | 139 | 91 | 136 | 190 |
| 60 -64 | 129 | 81 | 126 | 182 |
| 65 - 69 | 119 | 70 | 116 | 171 |
| 70 -74 | 110 | 62 | 107 | 161 |
| 75 - 79 | 101 | 52 | 97 | 152 |
| 80 - 84 | 93 | 44 | 88 | 144 |
| 85 - 89 | 82 | 35 | 78 | 135 |
| 90 -95 | 76 | 25 | 71 | 131 |

After tube voltage calibration, scanner model calibration was performed and the respective values for each scanner models are listed in **Table 2b**. The correction factor to calibrate individual scanner models to the selected reference scanner was less than 5% for most (41 of 50) scanner models and less than 10% for 47 or the 50 scanner models **(Table 2b).** This suggests that scanner model specific calibration is less critical than correction for tube kilovoltage difference.

### Normative Values

Osteoporosis prevalence among women aged 60–69 undergoing DXA screening at our institution (n= 95,465) was 23.74%. Analyzing a subset of these patients (n = 6,373) who underwent a CT scan within 12 months of their DXA screening, osteoporosis prevalence was near identical (23.65%). This further supports the hypothesis that a general indication for a clinical CT on average does not correlate with a higher or lower likelihood of osteoporosis in a significant way due to the wide variety of indications for a CT, most of which are not related to higher or lower bone density. Therefore, our study population was representative of the general population with regards to their statistical bone density distribution.

Attenuation values of trabecular bone of thoracic and lumbar vertebra stratified by patient age and sex were calculated. The results for the L1 vertebra by age and sex are provided in **Tables 3a and b** and **Figure 3**.

**Figure 3.**
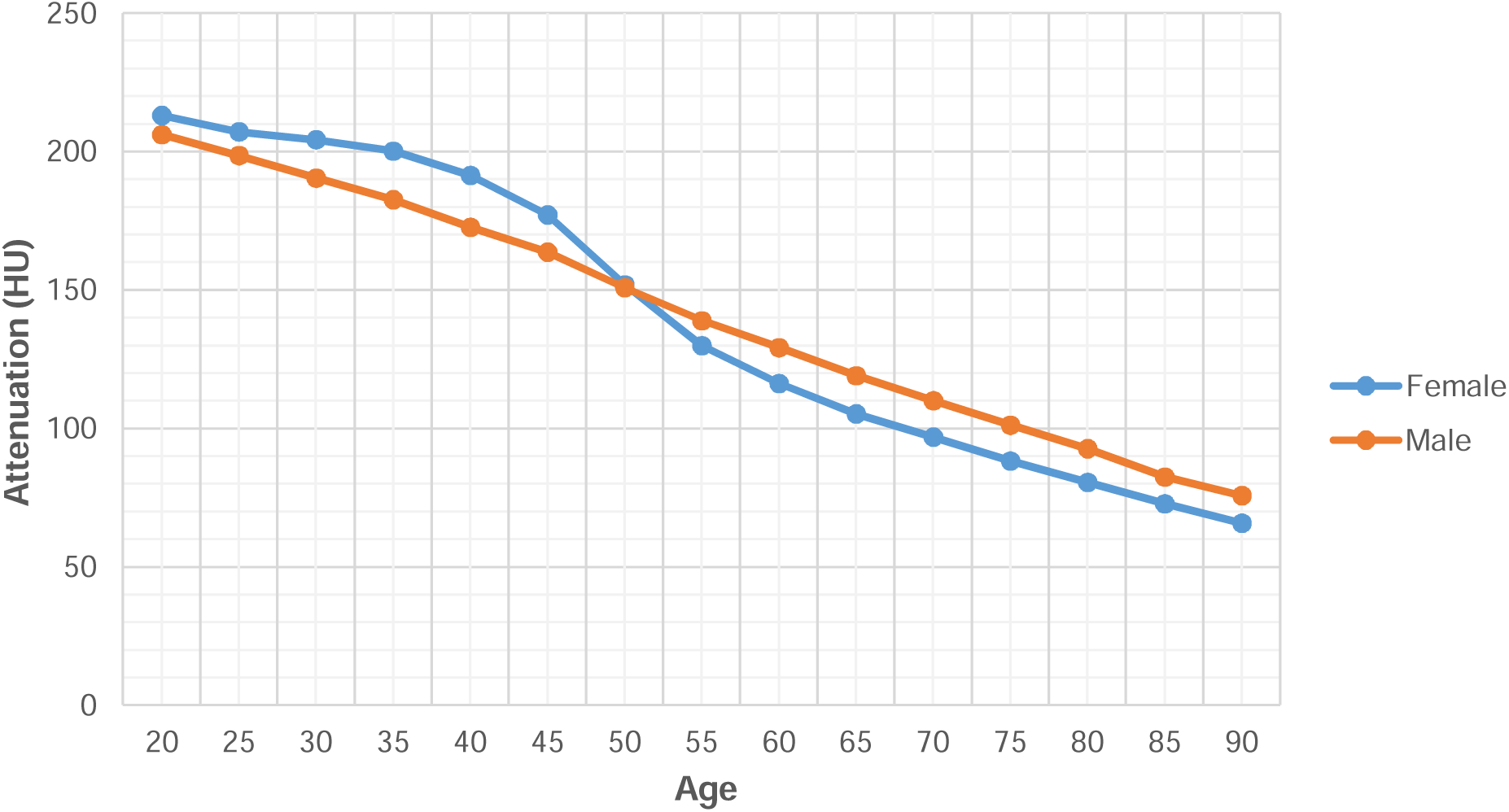
Mean trabecular attenuation in Hounsfield Units (HU) of L1 by sex and age. The decrease of trabecular attenuation in men was almost linear with age while in women there was a steeper drop around menopause. In all figures showing age, data points are aggregated in 5-year intervals and are labeled with the lower bound of the interval, e.g. the data point labeled 50 includes 50 – 54 years.

The mean trabecular attenuation decreased with increasing age. In younger patients (premenopausal, < 50 years), the attenuation was higher in women than in men (mean attenuation of L1 in 30–35-year-old women versus 30-35-year-old men: 204±42 HU versus 190±41 HU, p<.001), but lower in older (postmenopausal, >50 years) women than in men (mean attenuation of L1 in 65–70-year-old women versus 65-70-year-old men: 105±44 HU versus 119±44 HU, p<.001) **(Figure 3)**. The decrease of trabecular attenuation in men was almost linear with age while in women there was a steeper drop around menopause **(Figure 3).**

The attenuation measurements differed amongst the vertebrae. In general, the attenuation was lower in the more caudal vertebrae. It was highest in T1, and lowest in L3, though, in young men the difference between L1 to L4 was minimal. The age-related reductions in attenuation were greatest in the more caudal vertebrae, ranging from 2.3 HU/year in the T1 vertebra to 2.9 HU/year in the L5 vertebra in women, while the mean attenuation decreased from 1.2 HU/year (T1) and 2.1 HU/year (L5) in men (**Figure 4)**.

**Figure 4.**
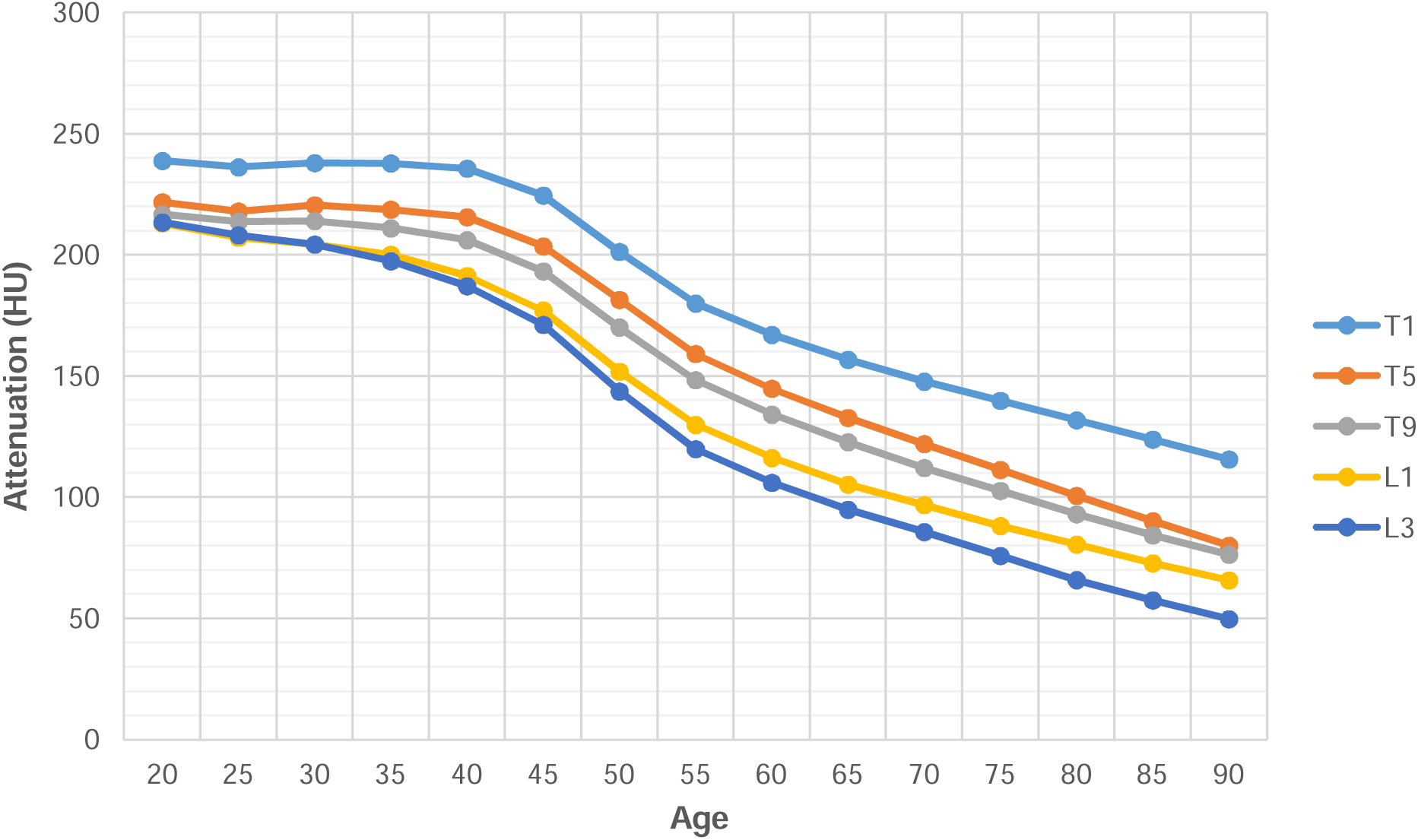
Age-related mean trabecular attenuation of selected thoracic and lumbar vertebra in Hounsfield Units (HU) in females. Attenuation differed amongst the vertebrae with lower attenuation in the more caudal vertebrae which also showed greater age-related reductions in attenuation.

**Figure 5a.**
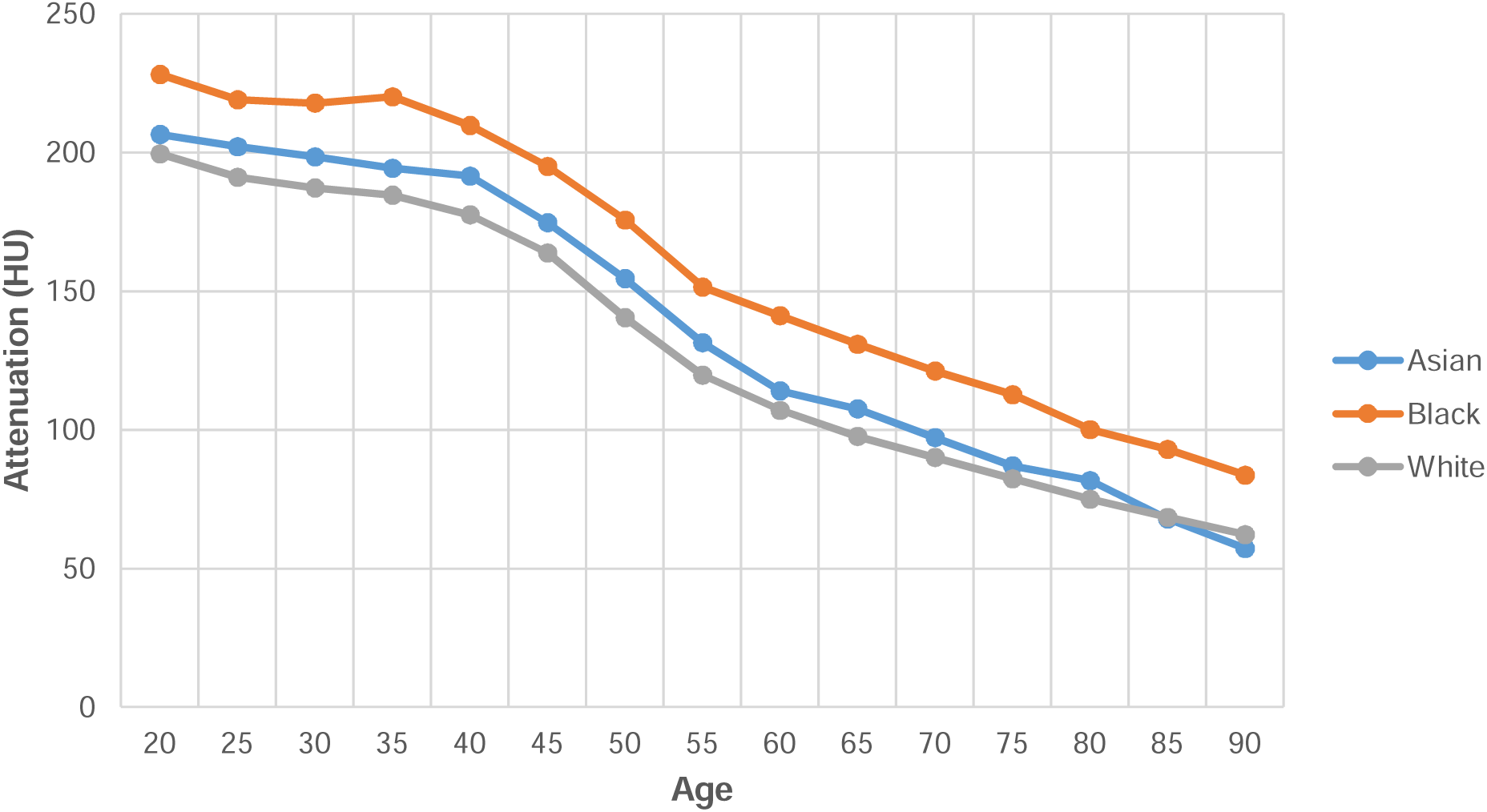
Mean trabecular attenuation in Hounsfield Units (HU) of the L1 vertebra in White, Black, and Asian females. Trabecular attenuation was highest in Blacks followed by Asians and lowest in Whites.

**Figure 5b.**
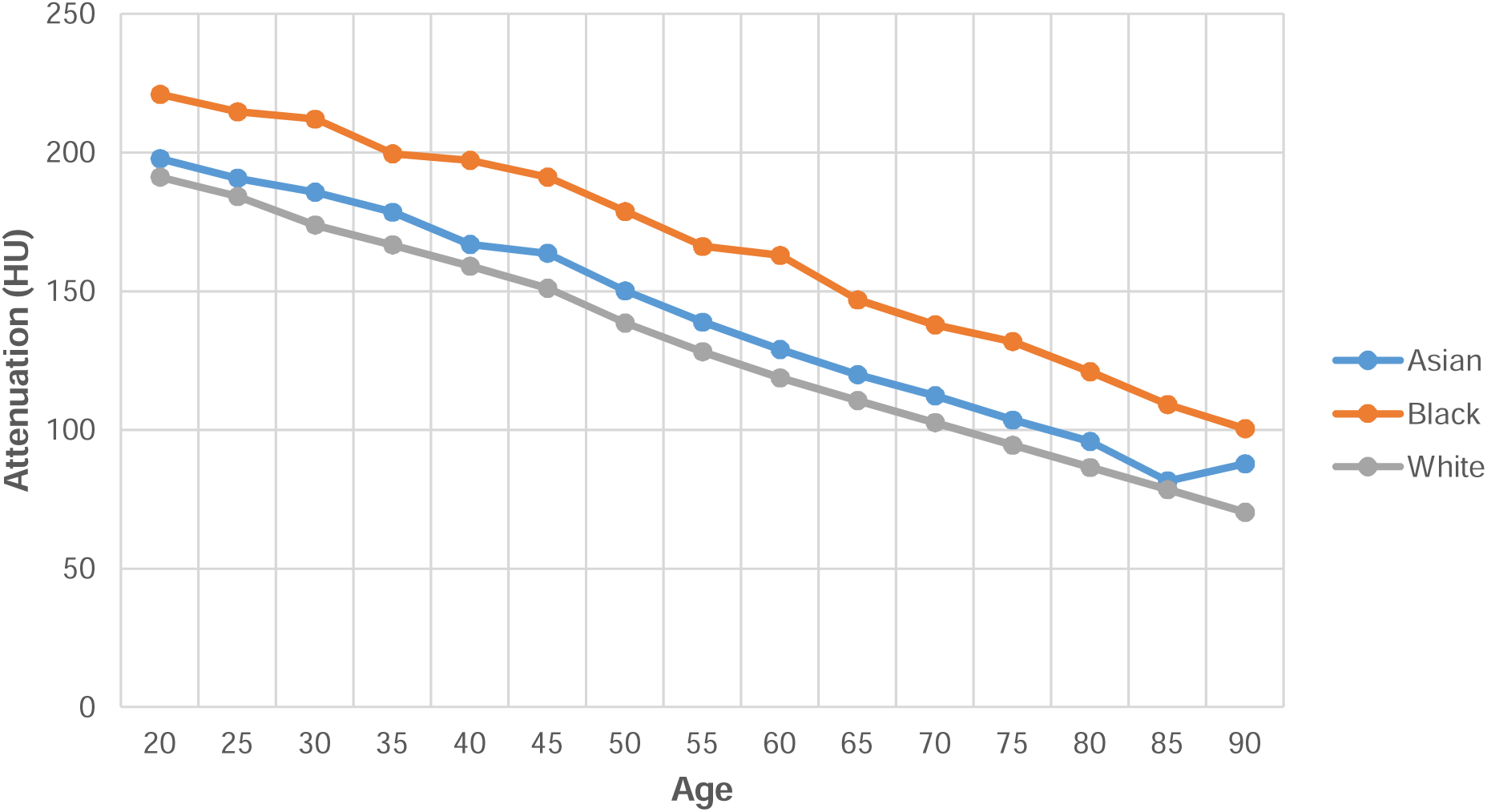
Mean trabecular attenuation in Hounsfield Units (HU) of the L1 vertebra in White, Black, and Asian males. Trabecular attenuation was highest in Blacks followed by Asians and lowest in Whites.

**Figure 6a.**
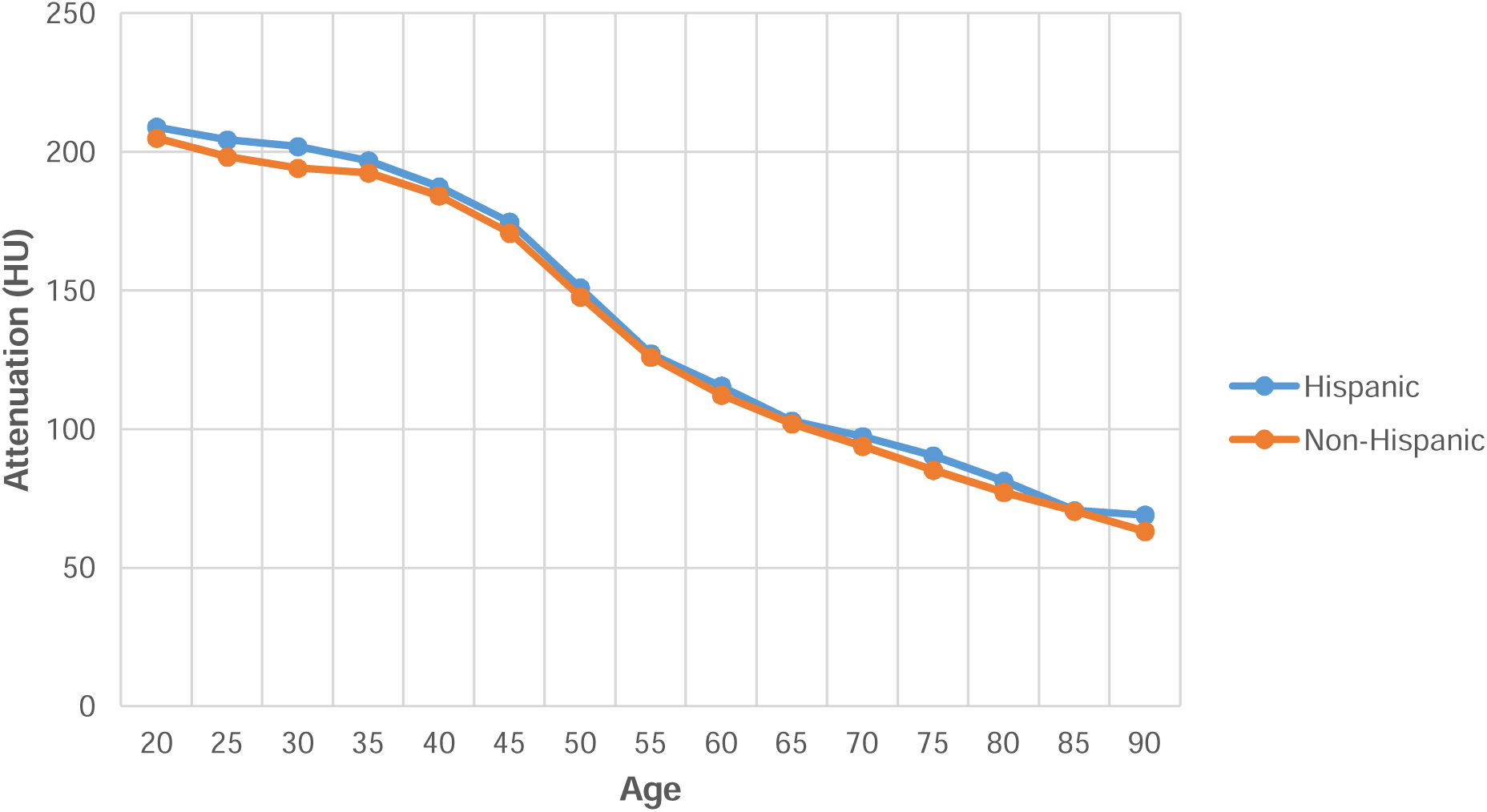
Mean trabecular attenuation in Hounsfield Units (HU) of the L1 vertebra in Hispanic and Non-Hispanic women. There was no statistically significant difference in trabecular attenuation in Hispanic and non-Hispanic women.

**Figure 6b.**
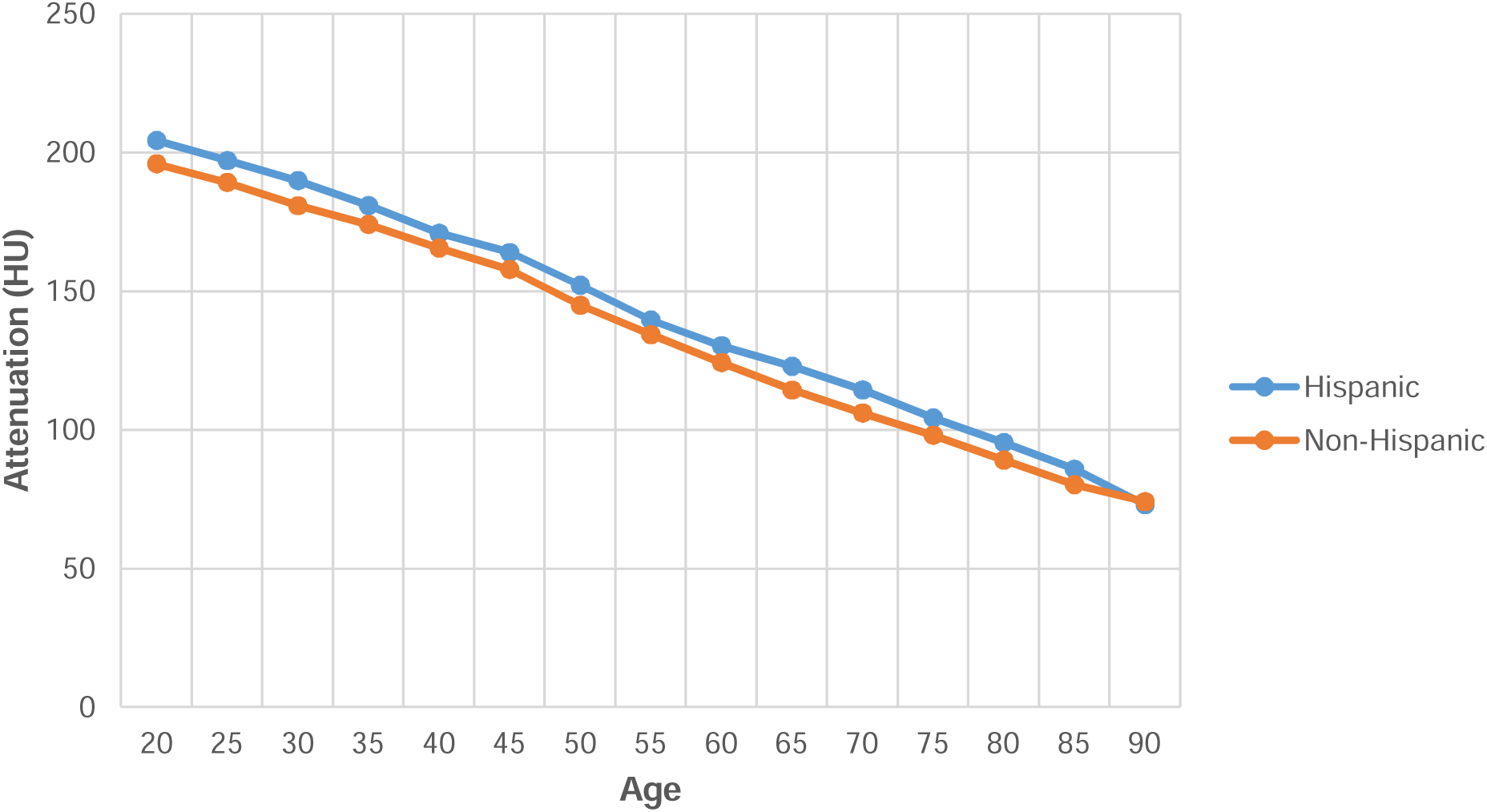
Mean trabecular attenuation in Hounsfield Units (HU) of the L1 vertebra in Hispanic and non-Hispanic men. Hispanic men had higher trabecular attenuation than non-Hispanic men.

The spread of trabecular attenuation was wide in both female and male patients, as shown as 10^th^ and 90^th^ percentile in **Tables 3a and b**. **Table 4** shows the 22^nd^ percentile and the 65^th^ percentile of the attenuation in each vertebra in females 60-69 years of age. As the prevalence of osteoporosis and osteopenia in this age group is estimated to be 22% and 65%, respectively (25), the values in **Table 4** can be used in men and women as thresholds to classify osteoporosis and osteopenia that would result in a similar incidence (22, 25). For L1, that threshold was 80 HU for osteoporosis and 117 HU for osteopenia.

**Table 4.** 22^nd^ and 65^th^ percentile of trabecular attenuation in HU in female patients 60-69y old.

| **Vertebra** | **22<sup>nd</sup> percentile Attenuation (HU)**<br>**osteoporosis** | **65<sup>th</sup> percentile Attenuation (HU)**<br>**osteopenia** |
| --- | --- | --- |
| T1 | 124 | 170 |
| T2 | 117 | 162 |
| T3 | 110 | 156 |
| T4 | 106 | 151 |
| T5 | 103 | 147 |
| T6 | 99 | 142 |
| T7 | 95 | 137 |
| T8 | 93 | 134 |
| T9 | 95 | 136 |
| T10 | 99 | 140 |
| T11 | 96 | 136 |
| T12 | 87 | 125 |
| L1 | 80 | 117 |
| L2 | 73 | 112 |
| L3 | 69 | 106 |
| L4 | 70 | 108 |
| L5 | 79 | 119 |

### Normative Values by Race and Ethnicity

Analyses were performed by self-reported race and ethnicity. The three largest groups were White (55.5%), Black (8.5%) and Asian (4.9%). In 31% no information on race was available **(Table 1).** The characteristics of age-related bone loss were similar in all three groups in women and men.

Trabecular attenuation was highest in Blacks followed by Asians and lowest in Whites (mean attenuation of L1 in 65–70-year-old Black, Asian, and White women: 131±44 HU, 108±42 HU, 98±32 HU, p<.001, and in 65–70-year-old Black, Asian, and White men: 146±47 HU, 120±41 HU, 111±37 HU, p<.001) **(Figures 5a and b).** In our cohort, 7.8% were Hispanic and 53.8% were non-Hispanic, while in 38.4% no data on ethnicity were available **(Table 1).** The characteristics of age-related bone loss were similar in both groups in women and men. There was no statistically significant difference in trabecular attenuation in Hispanic and non-Hispanic women (mean attenuation of L1 in 65–70-year-old Hispanic and non-Hispanic women:103±34 HU versus 102±37 HU, p=.36.) **(Figure 6a)**, while Hispanic men had higher trabecular attenuation than non-Hispanic men (mean attenuation of L1 in 65–70-year-old Hispanic and non-Hispanic men: 123±50 HU versus 114±40 HU, p<.001) **(Figure 6b).**

## Discussion

Osteoporosis is underdiagnosed and undertreated, and many patients at risk for osteoporosis are not referred for DXA screening, especially the underserved (2, 4–6). Our study showed that opportunistic osteoporosis screening, which leverages existing CT scans performed for other clinical purposes, can address these disparities and identify patients with low BMD who may be at risk for fractures. In our study of 538,946 CT examinations performed in 283,499 patients, an automated deep learning-based algorithm was able to accurately place a three-dimensional region of interest within thoracic and lumbar vertebra to measure trabecular attenuation using different scanner models and imaging protocols. There are technical challenges associated with opportunistic osteoporosis screening, such as variations in scan protocols or differences between scanner models (16, 17). This is mainly due to different compositions of the x-ray spectra with different tubes and different voltages, and the dependency of absorption rates of different materials on photon energy. These factors can lead to imprecise measurements of trabecular attenuation, potentially resulting in the overdiagnosis or underdiagnosis of osteoporosis (16). Phantom measurements, performed either synchronous with the clinical scan or asynchronous, can be used to determine the function that maps attenuation values obtained at one kVp setting on one scanner model to those measured at another (26, 27). However, phantom measurements are not practical for opportunistic screening applications and are generally not available for retrospective analysis. We therefore developed a novel statistical method of calibrating for different scan protocols and different scanner models. We found that tube voltage had a larger influence on attenuation values (23%) than scanner model (<10%), suggesting that scanner model specific calibration is less critical than correction for voltage difference.

The correction factors for scanner model and tube voltage from this study can be used by others to analyze retrospective and prospective datasets that were obtained using different scan protocols and scanner models.

Another technical challenge of opportunistic osteoporosis screening is accurate placement of the ROI within trabecular bone. Differences in slice selection or ROI placement can lead to substantial variations in results. In a study by Jang et al (14), automated ROI placement in the L1 vertebral body was on average 21 HU higher than manual placement, highlighting the need for accurate and reproducible ROI placement. While 3D ROIs, such as spheres or cylinders, provide more robust statistical measures by sampling multiple slices and more voxels, they are rarely used in clinical practice due to PACS limitations and the increased complexity of user interaction. We developed a novel method of automatically placing a 3D spherical ROI within trabecular bone which uses the lowest areas of trabecular attenuation, thereby avoiding bone islands or other lesions that can artifactually increase bone density.

Our measurements were lower than previously reported thresholds (14, 28). Compared to the 2D method described by Jang et al (14), the values using our method were a mean of 35 HU and median 26 HU lower, when averaged over all subjects. Thus, our threshold of 80 HU for diagnosing osteoporosis in a 3D ROI is comparable to a threshold of 100-115 HU using a 2D method reported in previous studies.

Our study showed that the trabecular attenuation differed in different vertebrae, indicating that specific thresholds should be used for each vertebra as indicators for osteoporosis, and provide reference values for each vertebra. Of note, age-related bone loss was greatest in the lumbar spine, suggesting that using lumbar vertebrae may be more sensitive to detect bone loss. Most prior studies limited evaluation to a single vertebra, such as the L1 (14, 29). Our automated method of placing 3D-ROIs makes it feasible to measure trabecular attenuation in all vertebrae in the field of view. The automatic generation of our attenuation measurements will be an important contributing factor to its widespread adoption, as it will decrease potential resistance from radiologists who could find the task of manually performing these measurements on eligible CT examinations burdensome.

One major obstacle to using opportunistic osteoporosis screening is the lack of normative data in diverse populations. Our study examining 538,946 CTs in 283,499 patients from different races and ethnicities is the largest to date. Our values show higher mean trabecular attenuation in Blacks compared to Whites and Asians in both sexes, which is consistent with findings previously reported in literature (30). We also show higher trabecular attenuation in Asians compared to Whites. Hispanic men had higher trabecular attenuation compared to non-Hispanics, while there was no significant difference in women. Clinical implications of opportunistic osteoporosis screening and established CT based normative values include an increase in the number of patients who will get screened for osteoporosis, and referred for DXA, enabling timely intervention, thereby preventing fractures, reducing morbidity, costs and improving overall patient outcomes (31–33).

Our study had several limitations. First, the generalizability of our study results may be limited as the data used to create the normative values were taken from facilities on the Northeastern United States. The inclusion of 283,499 patients should help negate the impact of this issue. Second, we used a 3D-ROI that identified the lowest attenuation within a vertebral body, and our attenuation values were a mean of 35 and a median of 26 HU lower than reported values that were obtained using a 2D methods. Therefore, our thresholds should not be used with 2D-ROIs or centrally placed 3D ROIs without adjustments. Third, while we implemented an automated method to determine and exclude incorrect segmentations, e.g. due to metallic implants or deformed vertebrae, our method might have not excluded all abnormal vertebrae. Fourth, we included patients without taking into account their past medical histories and medications which could potentially impact some of the measurements used to create our normative values. However, we modeled our methods based on general prevalence in the population, as it is the case for DXA thresholds, and therefore it is justified not to exclude patients on bone-active medications (20, 34). We showed that the prevalence of osteoporosis in a DXA screening cohort in our institution was near identical to the subset of those patients that also had a CT, suggesting that having an indication for CT in general is not a predictor for lower or higher bone mass. Fifth we did not validate our method against DXA-based diagnosis.

In conclusion, we developed a reproducible and robust deep learning-based method using a CNN that can automatically and accurately assess trabecular attenuation of the thoracic and lumbar spine on CT scans irrespective of the protocol or scanner model used during imaging acquisition. Age, sex, race and ethnicity stratified normative values were also established that can be used to diagnose osteoporosis.

These are important steps towards the widespread adoption of opportunistic CT imaging for this patient population, which can lead to improved patient outcomes through opportunistic osteoporosis screening and decreased overall healthcare costs.

## Abbreviations

CT: Computed Tomography
AI: Artificial Intelligence
DXA: dual-energy X-ray absorptiometry
ROI: region of interest
HU: Hounsfield Units
CNN: convolutional neural network

## Data Availability

All data produced in the present work are contained in the manuscript

