## Supplement for "Deep Learning-Based Opportunistic CT Osteoporosis Screening and Establishment of Normative Values"

**Supplement 1-**

**Automated Consistency Checks**

Anatomic variations or pathology, such as compression fractures, sclerotic or lytic lesions, or degenerative changes, as well as image quality problems, e.g. metal artifacts due to implanted hardware, can cause a segmentation to fail. In the clinical application on individual patients that is very easy to detect in an appropriate rendering of the segmentation, but for an automated analysis of many patients an automated method was required that can detect segmentation errors. We therefore performed the following checks on the segmentation. If one of the checks failed, the series was excluded from analysis:

1. A segmented vertebral body, that is not touching the boundaries of the field of view (typically the most cranial and most caudal axial slice) must be a single connected component.
2. We identify the most cranial and most caudal vertebral body that is present in the segmentation and require all anatomically in-between vertebrae to be present as well. For example, if the T12 and the L5 are present, then L1, L2, L3, L4 must be present as well.
3. The vertebrae must be geometrically in the correct order. For that we calculate the dot product of the vector connecting the centers of mass of one pair of anatomically adjacent vertebra with the vector connecting the centers of the next pair and require it to be >0. E.g. V(L1-L2) * V(L2-L3) > 0, or in other words the angle between those vectors must be > 90 degrees.
4. The distance between the centers of mass of adjacent pairs of vertebrae must not differ too much. This would be an indication for the segmentation having incorrectly fused to adjacent vertebra or split on vertebra, or very severely degenerated anatomy. We require the ratio of the vectors mentioned above to be between 0.67 and 1.5.
5. We use a simple metal detection, by applying a 3x3 averaging filter and then counting the pixels with >2200 HU. If the sum of such pixels in all axial slices that any individual vertebral body spans is >200 then we exclude the entire series. Note that even if the metal pixels are not contained within the segmented vertebra, we exclude the series, because significant amounts of metal can create artifacts, e.g. darker streaks, across the entire image.

It was validated that these Consistency Checks can catch the vast majority of incorrect segmentations by manually reviewing 128 randomly selected data sets that have passed consistency checks containing a total of 1496 lumbar and thoracic vertebrae. Three radiologists reviewed all 128 data sets to confirm that the spherical ROI is in fact placed correctly located inside the trabecular bone and in the correctly labeled vertebrae. In only one vertebral body in one scan, the ROI was incorrectly placed, namely inside an osteophyte adjacent to the vertebral body, that was incorrectly segmented as part of the vertebral body. The ROIs in all other 1495 vertebral bodies were correctly placed according to the three radiologists. This was determined to be sufficiently accurate to result in accurate and meaningful mean and percentile values for a large cohort.

**Supplemental Figure 1** Illustration of typical segmentation errors. detected by automated consistency checks.

**A**: The T11 vertebral body is labeled as T11 and T12 (white box) in this patient without a 12th rib.

E (C): consistency error

**B**: Metal artifact from fusion hardware.

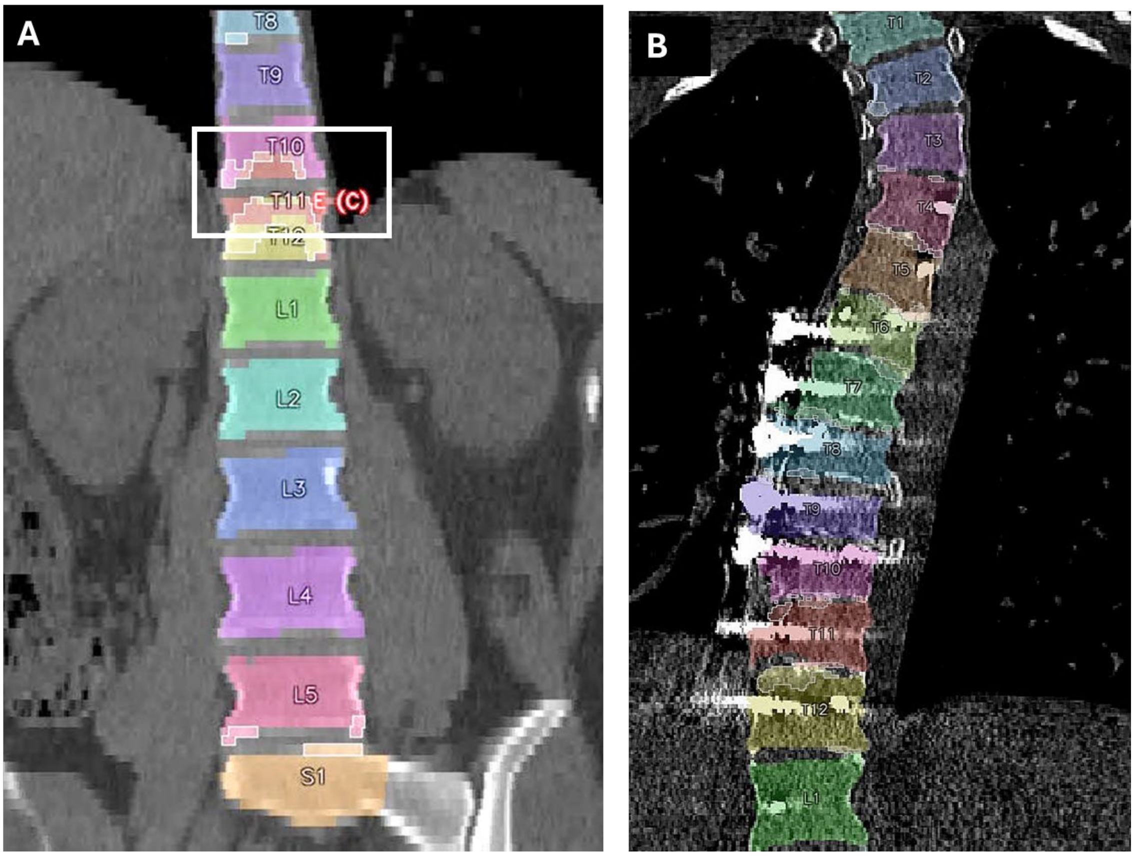
A
T8
T9
T10
T11
E (C)
T12
L1
L2
L3
L4
L5
S1
B
T1
T2
T3
T4
T5
T6
T7
T8
T9
T10
T11
T12
L1

**Supplement 2**

**Phantomless statistical calibration for kVp and scanner models**

While Hounsfield Units (HU) are calibrated for air and water ($HU_{Air}$ = -1000, $HU_{Water}$ = 0), the attenuation values for other materials can differ across scans with different tube voltage (kVp), or if performed on different scanner models (1, 2).

We have developed a phantomless statistical approach for different tube voltages (kVp) and scanner models.

The general idea is as follows:

If we have two sufficiently large and comparable patient cohorts, we can assume that on average the bone mineral density of a specific vertebra is the same in both. If one cohort is scanned at e.g. 120kVp and the other at 90 kVp, we can compare the measured average attenuation to obtain our mapping function.

We can do that for each kVp, sex, age group, and vertebra in our cohort. For example, we determine the average attenuation in the L1 for all female patients 40-44 years scanned at 120 kVp (L1, F 40y, 120 kVp) and the same for all scans at 90 kVp (L1, F40 years, 90k Vp). Again, the 120kVp scans are (generally) in different patients but given the composition of the cohort is identical, on average their anatomical bone density is the same. We interpret these two averages as x and y coordinates of one data point.

We do the same for all age groups (in 5-year intervals), for all thoracic and lumbar vertebrae, and for female and male patients. This gives us over 300 data points. This is depicted for the example of 90 kVp and 120 kVp in the **Supplemental Figure 2**. We can then fit a mapping function to the data points. We have chosen a simple linear function with an offset of zero which we can obtain using simple linear regression.

The slope of that curve gives us our mapping function $HU_{120} = A_x * HU_{90}$ that maps an attenuation value measured at 90 kVp to what would have been measured at 120 kVp.

This allows us to calibrate all scans to 120 kVp, which we have chosen as reference standard, because it is the most common setting. As we show in **Table 2a** in the paper the correction can be significant, for example for 90 kVp it is 0.81, i.e. the correction is ca. 20%.

Due to different x-ray tube characteristics, filters and other scanner parameters different scanner models may have different x-ray spectra even at the same kVp. After calibrating for kVp we can therefore repeat the same method to map an individual scanner model A to another scanner model B.

We have done that for 50 scanner models for which we had a sufficient number of scans with results shown on **Table 2b** in the paper.

We have validated our method by scanning the FDA-approved QCTPro calibration phantom (Mindways Software, Inc., Austin, TX) at different voltages and in several different scanner models. The results are consistent with our statistically obtained results. Our results are also consistent with other published kVp results (1).

The difference between scanner models are smaller than the impact of tube voltage, mostly under ±5%. When not calibrated this could lead to a measurement error of 5-10 HU, while the difference between an osteoporotic and a normal bone is in the order of 100 HU.

We conclude that while correction for kVp and scanner model will produce more accurate results, our results suggest that even if calibration values are not available for a given scanner model, clinically reasonable results can likely be achieved even without scanner model calibration.

Limitations: Even in the same scanner, effects like beam hardening could impact measurement accuracy. Also, there could be variability between different scanners of the same type or even in one scanner over time. The standard CT calibration to Hounsfield units significantly reduces these effects. Nevertheless, either a phantom-based or a statistical calibration experiment should be performed for new scanner models and scanning protocols especially if these use techniques and technologies significantly different from the scanners used to obtain the reference data.

**Supplemental Figure 2** Illustration of regression analysis for mapping function from one specific scanning protocol (here 90 kVP) to 120kVP

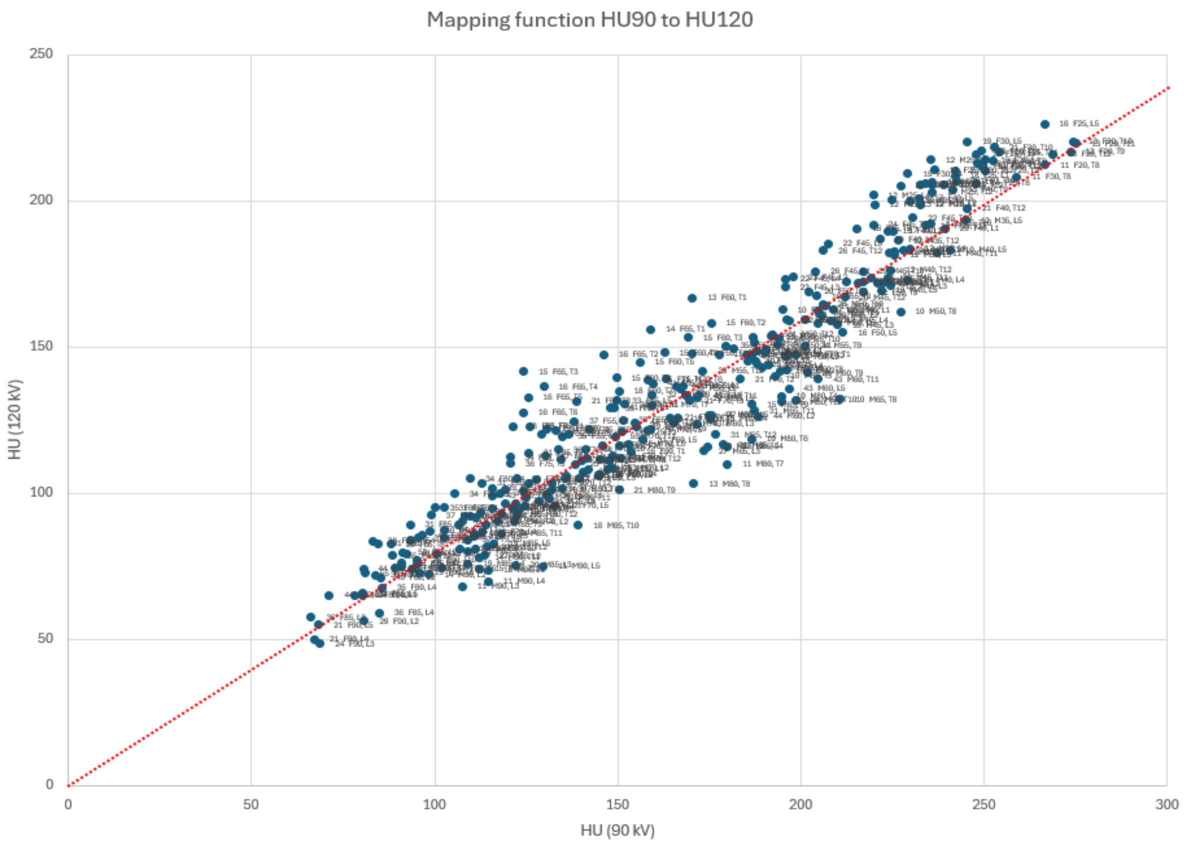


Such a regression analysis was performed for all kVP values 80kV, 90kV,100kV, 110kV ,120kV (identity) and 130kV. This figure illustrates the process for the example of 90 kVP. Each data point represents the average of all scans of a specific vertebra in a specific age group and sex measured at 120 kVp (y coordinate) and 90 kVp (x coordinate) respectively. The dotted line illustrates an offset free linear regression line.
